# LIRIC predicts Hepatocellular Carcinoma risk in the diverse U.S. population using routine clinical data

**DOI:** 10.1101/2024.05.28.24307949

**Authors:** Kai Jia, Bowen Gu, Pasapol Saowakon, Steven Kundrot, Matvey B. Palchuk, Jeff Warnick, Irving D. Kaplan, Martin Rinard, Limor Appelbaum

## Abstract

**Background and Aims:** Hepatocellular Carcinoma (HCC) is often diagnosed late, limiting curative treatment options. Conversely, early detection in cirrhotic patients through screening offers high cure rates but is underutilized and misses cases occurring in individuals without cirrhosis. We aimed to build, validate, and simulate the deployment of models for HCC risk stratification using routinely collected Electronic Health Record (EHR) data from a geographically and racially diverse U.S. population.

**Methods:** We developed Logistic Regression (LiricLR) and Neural Network (LiricNN) models for the general (GP) and cirrhosis populations utilizing EHR data from 46,79 HCC cases and 1,128,202 controls aged 40-100 years. Data was sourced from 64 Health Care Organizations (HCOs) from a federated network, spanning academic medical centers, community hospitals, and outpatient clinics nationwide. We evaluated model performance using AUC, calibration plots, and Geometric Mean of Overestimation (GMOE), the geometric mean of ratios of predicted to actual risks. External validation involved HCO location, race, and temporal factors. Simulated deployment assessed sensitivity, specificity, Positive Predictive Value, Number Needed to Screen for each risk threshold.

**Results:** LiricLR and LiricNN (GP) achieved test set AUCs of AUC=0.8968 (95% CI: 0.8925, 0.9010) and AUC=0.9254 (95% CI: 0.9218, 0.9289), respectively, leveraging 46 established (cirrhosis, hepatitis, diabetes) and novel (frequency of clinical encounters, platelet, albumin, aminotransferase values) features. Average external validation AUCs of LiricNN were 0.9274 (95% CI: 0.9239, 0.9308) for locations and 0.9284 (95% CI: 0.9247, 0.9320) for races. Average GMOEs were 0.887 (95% CI: 0.862-0.911). Simulated model deployment of LiricNN provides performance metrics across multiple risk thresholds.

**Conclusions:** Liric models utilize routine EHR data to accurately predict risk of HCC development. Their scalability, generalizability, and interpretability set the stage for future clinical deployment and the design of more effective screening programs.

**Lay Summary:** Hepatocellular Carcinoma (HCC), the most common liver cancer, is often diagnosed in late stages, limiting treatment options. Early detection through screening is essential for effective intervention and potential cure. However, current screening mostly targets patients with liver cirrhosis, many of whom do not get screened, while missing others who could develop HCC even without cirrhosis.

To improve screening, we created and tested Liric (LIver cancer RIsk Computation) models. These models use routine medical records from across the country to identify people at high risk of developing HCC.

Liric models have several benefits. Firstly, they can increase awareness among primary care physicians (PCPs) nationwide, improving the utilization of HCC screening. This is particularly crucial in areas with socio-demographic disparities, where access to specialist physicians may be limited. Additionally, Liric models can identify patients who would be missed by current screening guidelines, ensuring a more comprehensive approach to HCC detection.

Liric can be integrated into EHR systems to automatically generate a risk score from routinely collected patient data. This risk score can provide valuable information to physicians and caregivers, helping them make informed decisions about the need for HCC screening and can be used to develop cost-effective screening programs by identifying populations in which screening is effective.

**Graphical abstract:** 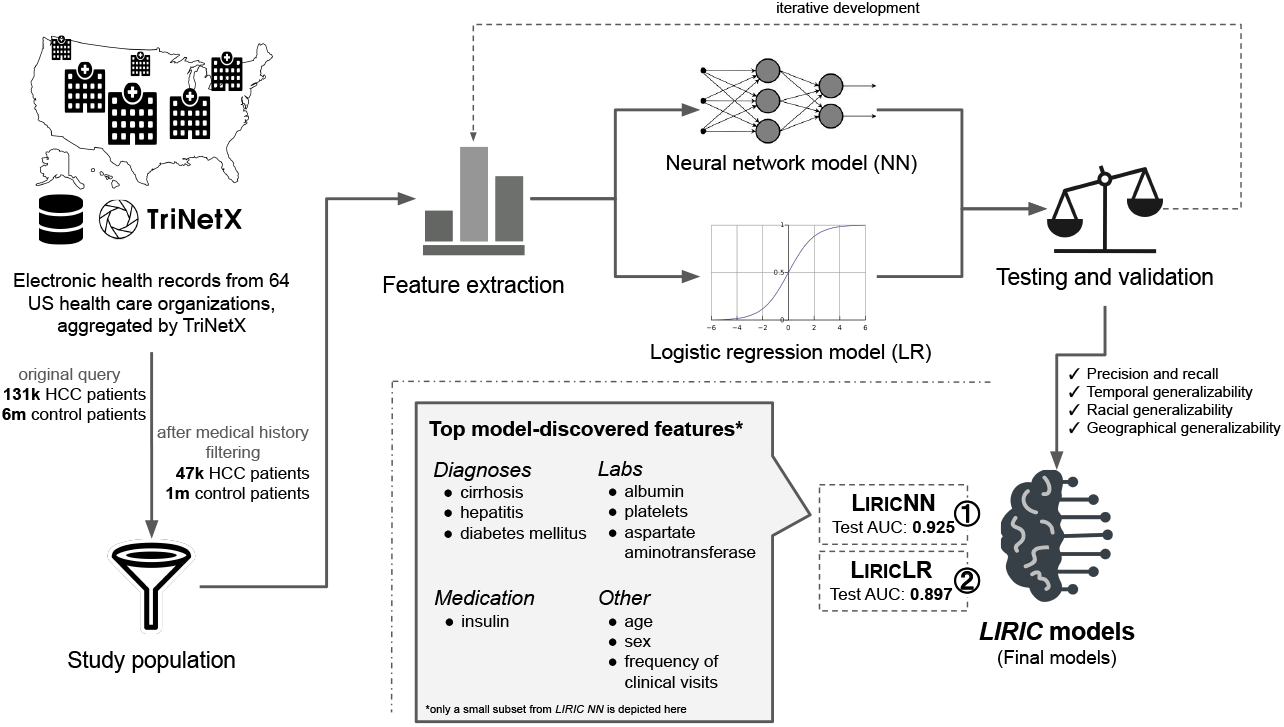

**Highlights:**

- Screening detects HCC early but is underutilized and misses cases without cirrhosis
- We developed, validated, and simulated deployment of Liric to identify individuals at high-risk for HCC
- Liric uses routinely collected clinical and lab data from a diverse US population
- Liric accurately predicts risk of HCC 6-36 months before it occurs
- Liric can assist PCPs in identifying individuals most in need of screening

**Impacts and implications:** Effective screening for hepatocellular carcinoma (HCC) is vital to achieve early detection and improved cure rates. However, the existing screening approach primarily targets patients with liver cirrhosis, and is both underutilized and fails to identify those without underlying cirrhosis.

Implementation of Liric models has the potential to enhance nationwide awareness among primary care physicians (PCPs), and improve screening utilization for hepatocellular carcinoma (HCC), particularly in regions characterized by socio-demographic disparities. Furthermore, these models can help identify patients who are currently overlooked by existing screening guidelines and aid in the development of new, more effective guidelines.

Integration of Liric models into EHR systems via a federated network would enable automatic generation of risk scores using unfiltered patient data. This approach could more accurately identify at-risk patients, providing valuable information to caregivers for HCC screening.

## Introduction

Hepatocellular Carcinoma (HCC) is commonly detected in its advanced stages, with a 5-year survival rate of 2.5% for metastatic disease [1]. However, diagnosis of early-stage cancer can lead to cure by surgical resection, ablation, or transplantation [2], significantly enhancing survival rates [3], with up to 70% of individuals achieving a five-year survival [4]. Screening for HCC in high-risk patients using liver Ultrasound (US) and alpha-fetoprotein is well-established, leading to detection of early-stage disease and high cure rates [5, 6], as well as decreased costs to the healthcare system [7, 8].

Current screening guidelines consider high-risk patients to be those with underlying cirrhosis from any cause, including viral (HBV, HCV) and non-viral (alcohol, NASH), or chronic hepatitis B infection with certain high-risk features [4, 9, 10, 11, 12]. However, about 13% of individuals that develop HCC do not meet screening eligibility criteria because they do not have underlying cirrhosis or have undiagnosed cirrhosis [13]. Moreover, screening is underutilized, with less than 20% of eligible patients undergoing screening, due to a combination of physician related factors, such as lack of primary care physician (PCP) awareness, as well as patient socio-demographic factors. Patients seen by specialists, usually in higher-income urban areas, are much more likely to be referred to and comply with screening [5, 14, 15, 16, 17].

Risk prediction models using patient data present a noninvasive, relatively inexpensive means of focusing screening efforts on a targeted population to identify individuals at higher risk for HCC. However, existing models have focused only on the cirrhotic population [18, 19], overlooking HCC cases without underlying cirrhosis. Others have developed general population models [20, 21, 22, 23], but these were developed on small patient numbers using population data derived from a single Asian country (Korea, Japan, Taiwan), and lack validation on an independent population [20, 21, 22, 23]. In one model for which external validation was reported, it was tested on an independent population but from the same country (Korea), limiting its generalizability [24]. Therefore, these models may not be transferable to other diverse populations in different geographic regions around the globe. Moreover, most existing population models use simple statistical regression methods and a predefined set of known features [21, 22, 24] and lack discovery of novel features which could improve predictions, as well as lead to causal association studies. Those that do use machine learning typically rely on a predefined set of known features [23]. Importantly, published models lack an obvious path to integration within EHR systems. Such a roadmap must enable simple and automated model deployment, which is a critical step for clinical implementation and physician adoption of models. Lack of such a roadmap presents a significant future hurdle obstructing real-world clinical use [25].

Our aim was to develop, validate, and simulate the deployment of accurate, generalizable, and interpretable HCC risk prediction models for the general population and for patients with liver cirrhosis, using routinely available clinical and lab features derived from patient EHRs within a federated network platform. Since we used data from health care organizations across the US, including about 47,000 HCC cases and 1.1 million controls, these models can potentially be used in primary care settings to identify individuals at elevated risk for HCC from diverse races and geographic locations.

## Materials and methods

We used the TRIPOD (Transparent Reporting of a multivariable prediction model for Individual Prediction or Diagnosis) guidelines for conducting and reporting on model development and validation [26]

All EHR data were obtained through the federated global health research network, TriNetX, and de-identified by TriNetX. We accessed the data under a no-cost collaboration agreement. Based on a determination by the Western IRB, studies using TriNetX data are not considered to be human subject research, and are therefore exempt from IRB review.

We adopted a methodology consistent with our prior study [27], with modifications tailored to the specific objectives of the current study. Briefly, we trained two classes of models: Neural Networks (NN) and Logistic Regression (LR), employed our feature selection algorithms to improve interpretability, conducted three types of internal-external validation, and simulated model deployment in a prospective setup. These methods were replicated with appropriate adjustments to suit the nuances of the present research.

### Study design

We used multi-institutional unfiltered EHR data to develop two types of models for predicting HCC risk: A general population model, and a population model that only includes individuals with liver cirrhosis. In this retrospective observational study, two study designs were utilized: a case-control design for developing the models, and a cohort design for simulating their deployment.

### Data source and setting

TriNetX is a federated global health research network that specializes in data collection and distribution [28]. The TriNetX database encompasses data from a diverse array of healthcare organizations (HCOs), spanning academic medical centers, community hospitals, and outpatient clinics.

We employed de-identified historical EHR data from 64 HCOs from throughout the United States, with each HCO contributing about 13 years of data on average. The data includes structured EHR fields such as demographics, date-indexed encounters, diagnoses, procedures, labs, and medications. It also contains facts and narratives extracted from unstructured text through the application of Natural Language Processing (NLP). TriNetX standardizes data from different HCOs according to the TriNetX standard data model with a uniform set of curated terminologies. To ensure data quality, TriNetX also has systems in place to detect anomalies and outliers XXX - what do they do about these anomalies and outliers XXXX.

Our study involved four groups: general hepatocellular carcinoma (HCC) case groups, hepatocellular carcinoma (HCC) case groups with cirrhosis, and their corresponding control groups. The general HCC cases and control group data was queried from TriNetX on Jan 26th, 2024. The HCC with cirrhosis cases and control group data was queried from TriNetX on Apr. 10th, 2024.

### Study population for development of the general population models

For the general HCC case group, we queried the TriNetX database for patients aged 40 or more with an ICD-10 code of 22.0, for a total of n=130,907 patients. We then excluded those diagnosed with HCC before turning 40 (n=4,116), those with no medical history 6 months prior to their HCC diagnosis (n=34,789), and those with records 2 months after their death record (n=1,032). The remaining n=46,679 patients were enrolled in the HCC group.

To obtain the general HCC control group, we queried the TriNetX database for patients aged 40 or more without an ICD-10 code of 22.0 or an ICD-9 code of 155.0, a total of n=59,823,821 patients. Then, a number (n=6,000,004) of patients were randomly selected. Among these patients, we excluded those with an HCC tumor registry (n=180), those whose last entry was before age 35.5 (n=821,083), those with less than 90 days of medical history (n=1,578,489), and those with records 2 months after their death record (n=35,238). The remaining n=1,128,202 patients were enrolled in the control group.

Cohort demographics, including sex, age, race, and HCO location, are shown in Table 1.

**Table 1:**
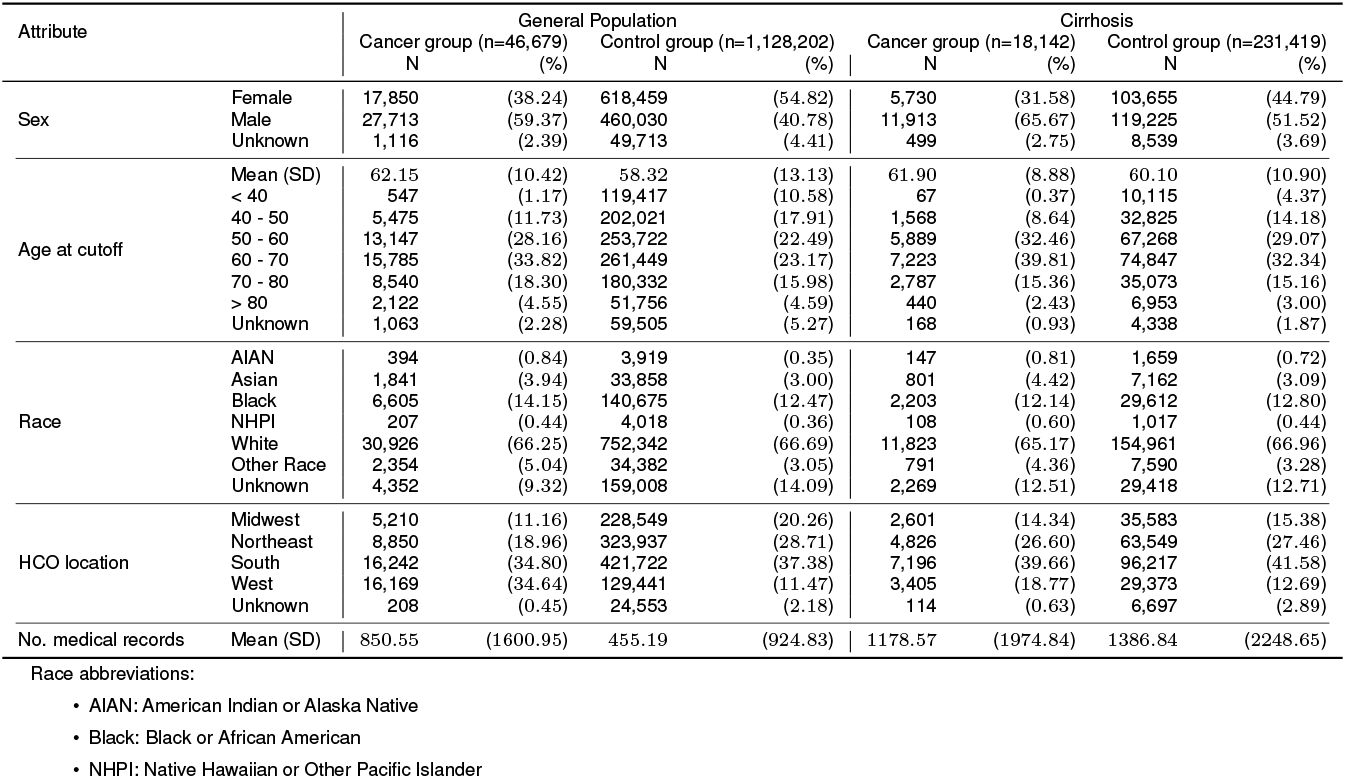
Demographics of our dataset.

### Study population for development of models for individuals with cirrhosis

For the HCC with cirrhosis case group, we queried the TriNetX database for patients aged 40 or more with an ICD10 code of 22.0 for HCC and an ICD-10 code of K70, K70.3, K70.30, K70.31, K74, K74.0, K74.6, K74.60, or K74.69 and an ICD-9 code of 571.2 and 571.5 for cirrhosis. Due to the cutoff date, we also require the diagnosis of HCC must be at least 7 months after the diagnosis of cirrhosis. This gives a total of n=44,680 patients. We then excluded those diagnosed with HCC before turning 40 (n=503), those with no medical history 6 months prior to their HCC diagnosis (n=16,357), those with records 2 months after their death record (n=262). The remaining n=18,142 patients were enrolled in the HCC group.

To obtain the HCC with cirrhosis control group, we queried the TriNetX database for patients aged 40 or more with an ICD-10 code of K70, K70.3, K70.30, K70.31, K74, K74.0, K74.6, K74.60 or K74.69 and without an ICD-10 code of 22.0. This gives a total of n=739,866 patients. We did not perform random sampling for this group as the total number of patients is already a small enough to be handled by our computing resources. Among these patients, we excluded those with an HCC tumor registry (n=168), those whose last entry was before age 35.5 (n=1,892), those with less than 90 days of medical history (n=139,378), and those with records 2 months after their death record (n=14,873). The remaining n=231,419 patients were enrolled in the control group.

Cohort demographics, including sex, age, race, and HCO location, are shown in Table 1.

### Model development

#### 1. Model classes

Two model classes were considered: neural networks (LiricNN) and logistic regression (LiricLR).

#### 2. Dataset partitioning

The dataset was randomly partitioned into training (75%), validation (10%), and test (15%) sets.

#### 3. Considerations on data inclusion

During training and testing, each patient was assigned a cutoff date. The model accessed data up to the cutoff date to predict HCC risk between 6 and 36 months after that date. For patients in the general HCC cohort, the cutoff dates were sampled uniformly between 6 and 36 months before the HCC diagnosis. For patients in the cirrhosis HCC cohort, the cutoff dates were sampled uniformly between 6 months before HCC diagnosis, up to 36 months before HCC diagnosis or 1 month after the cirrhosis diagnosis, whichever comes later. For patients in the control cohort, the cutoff dates were sampled according to the distribution of cutoff dates in the HCC cohort, but at least 1 month after cirrhosis diagnosis for the cirrhosis control cohort. We chose to use the time period, which excludes data from immediately 6 prior to HCC diagnosis, because this lag period seemed appropriate to account for a possible existing workup due to clinical suspicion for HCC. We chose to look back up to 3 years before diagnosis for the general and cirrhosis HCC cohorts, because we believe this to be the optimal time period for detecting early-stage resectable cancer with current screening modalities. We empirically defined a patient to have *sufficient medical history* if **(1)** the patient’s total number of diagnosis, medication, or lab entries in the 3-year window preceding their cutoff date was at least 16, **(2)** the patient’s medical entries prior to their cutoff date spanned at least 3 months, and **(3)** the patient’s were at least 37 years old (40 years old minus 36 months) on their cutoff date. Patients without sufficient medical history were excluded.

#### 4. Feature extraction

For each patient, we considered their EHR records up to their assigned cutoff date and turned them into a fixed number of features, which can be categorized into four classes: basic, diagnosis, medication, and lab.

Basic features encode patient demographics (age and sex) and clinical encounter frequencies (the numbers of *early* and *recent* EHR entries). Records from over 36 months before the cutoff date are considered early, and those from no more than 36 months before the cutoff date are considered recent.

Diagnosis, medication, and lab features share a similar structure. For each diagnosis, medication, or lab *entry type* (where an entry type might be a diagnosis of diabetes mellitus or a lab for creatinine in serum, for instance), they encode the existence, the frequency, and the time span that the entry type appears in a patient. Lab features additionally include lab results. Existence is encoded as a binary feature (0 or 1) that determines whether a given entry type ever occurs in a patient’s EHR (on or before the cutoff date). This accounts for the effect of the healthcare process on EHR data [29] and eliminates the need for data imputation as far as non-linear models such as LiricNN are concerned since they can make complicated decisions conditioned on whether a feature exists.

To limit the number of features, we discarded entry types that occurred in fewer than 1% of HCC cases in the training set. However, doing so still resulted in thousands of features, so we further employed *L*_0_ regularization on binary input mask [30] and iterative feature removal [27].

#### 5. Model calibration

We calibrated our models with a modified version of the Platt calibration [31] so that the model-estimated cancer risks align with the actual cancer risks. To do so, we modeled the logit of an individual’s person-year risk as *f*_*θ*_(*s*) = *θ*_1_ min(*s−s*_0_, 0) + *θ*_2_ max(*s−s*_0_, 0) + *θ*_3_, where *θ ∈* ℝ^3^, min(*θ*_1_, *θ*_2_) *>* 0, *s* is the model output score, and *s*_0_ is the median of all model output scores on the validation set. We then used linear regression to fit *θ* on the validation set, independently for each model.

#### 6. Performance measures

We trained our models nine times, each time with a different seed for data splitting and model initialization. We then measured the mean AUC and GMOE metrics achieved on the test sets.

To evaluate the risk calibration, we took models from the first run (of the nine runs) and generated calibration plots on the test set. For quantative calibration evaluation, we also calculated the Geometric Mean of Over Estimation (GMOE), the geometric mean of the ratios of predicted risks to the actual risks. A GMOE value close to 1 indicates that the predicted probabilities are well-calibrated.

#### 7. Selected model features

We present the features automatically selected by the LIRICNN model to interpret the model’s decision-making process [32]. The features are ranked by their univariate predictive power in LiricNN (from the first run), defined by the AUC achieved by using only that feature type (and zeroing out all other feature types).

### Internal-external validation

We performed external validation by partitioning large-scale federated network data based on certain attributes of interest [33]. Three attributes were considered: HCO geographic location, patient race, and training data time. For each attribute, we partitioned our dataset according to the attribute, trained models from scratch on one of the partitions, and tested them on the other partitions not used for training.

For validations on geographical location and patient race, we compared the AUCs achieved by external-validation models with those of control models. Both types of models were trained and tested with data of the same cases counts. The difference lay in the data splitting process, wherein external-validation models would be trained on data with a fixed attribute value (for example, location West) and tested on data with other attribute values (in the same example, all locations except West), while control models would be trained and tested on data that is split without consideration given to HCO locations (i.e., uniformly random splitting). Furthermore, we computed the gaps between validation-set and test-set AUCs for additional evaluation of generalizability. Please note that patients without HCO location or race specified were excluded.

For temporal validation, we trained models on data timestamped before certain *split dates*, chosen as the 50%, 60%, …, 90% percentile of diagnosis dates in the dataset. We ensured to use a uniform number of cases in training each model (by random sampling cases to match the target number of cases). All models obtained were tested on data timestamped after the 90% percentile (Oct 11, 2022 for the general HCC cohort and Feb 23, 2023 for the cirrhosis HCC cohort).

### Simulated deployment

To approximate the true performance of our models in a clinical setting, we conducted a simulated prospective study on the TriNetX database.

We trained models on data timestamped before 70% percentile of diagnosis dates (Feb 7, 2020 for the general HCC cohort and Dec 3, 2020 for the cirrhosis HCC cohort). Each individual in the test set was then periodically evaluated for HCC risk with those models, starting from Feb 7, 2020 (Dec 3, 2020 for the cirrhosis HCC cohort). Individuals identified as *high-risk* at any periodical test point would then be followed up to track their HCC incidence.

We did this for several high-risk thresholds, which were determined according to certain specificity levels on the validation set. We then computed sensitivity, specificity, and Positive Predictive Value (PPV).

We recognize the imbalanced nature of our dataset, which included all HCC cases but only a subset of control cases from the TriNetX database. To address this, for the general HCC cohort, we estimated the PPV values as though the models were tested on the entire TriNetX population, rather than using the actual PPVs from our limited test set (see the “PPV (TrxPop. Est.)” column of Table 2). For the cirrhosis HCC cohort, since we used all the cirrhosis cases found on TriNetX database, there is no need to perform and estimation of the PPV values. As a result, we report the actual PPV values in Table 3.

**Table 2:**
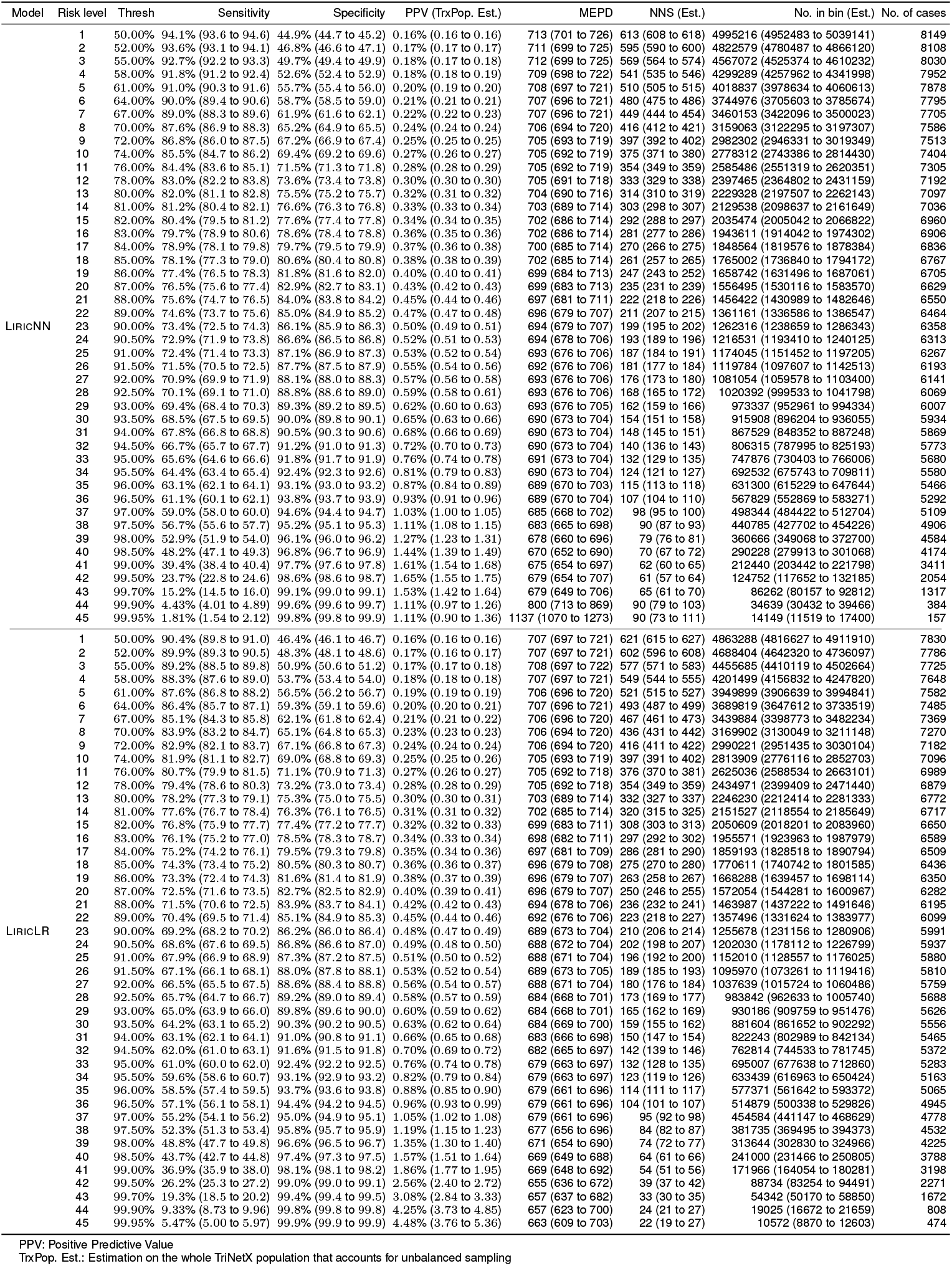
Simulated deployment results. Numbers in brackets are 95% CI (general population).

**Table 3:**
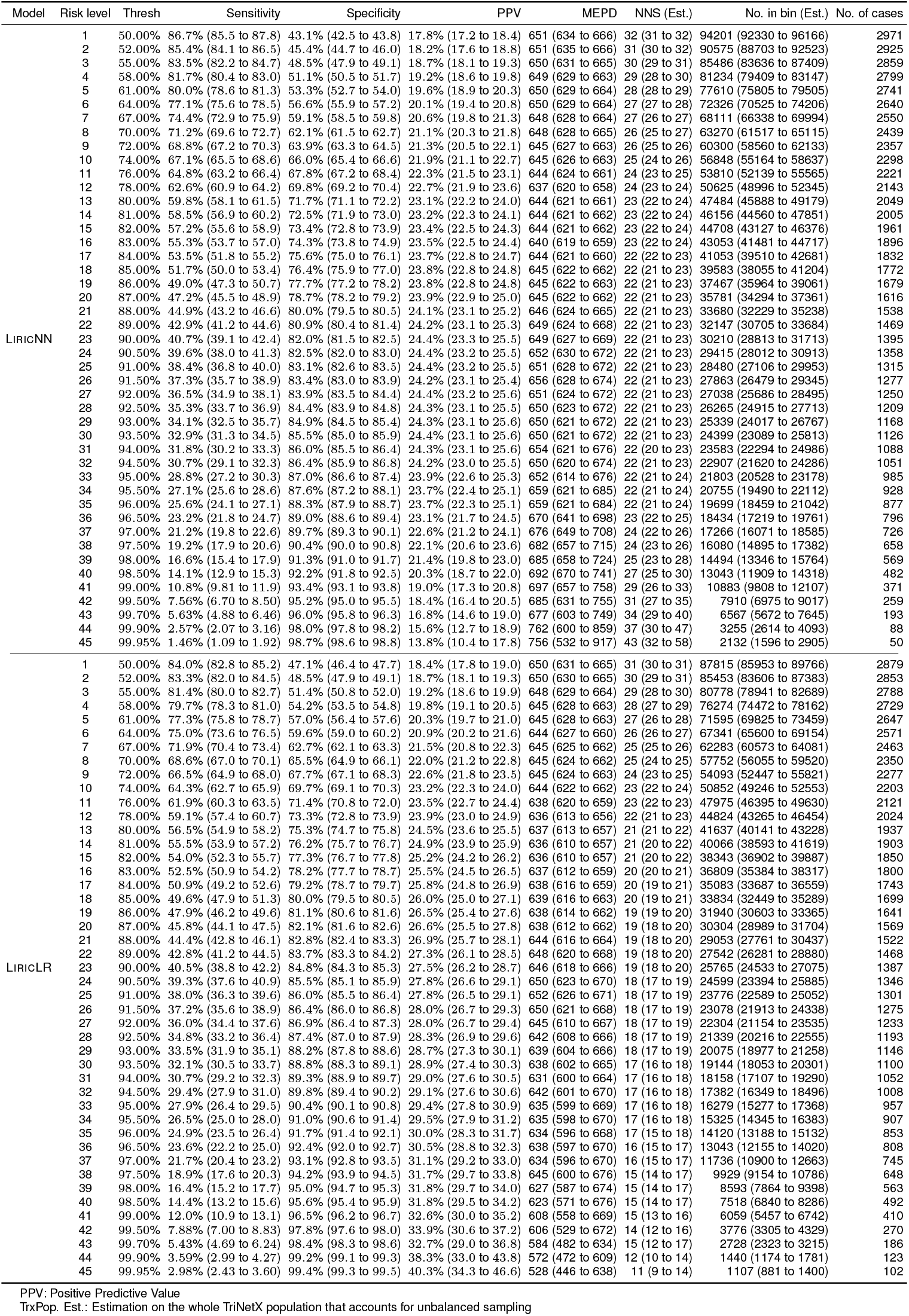
Simulated deployment results. Numbers in brackets are 95% CI (cirrhosis).

## Results

### Study population characteristics

For the general population, 66.25% of cases were of White race. Black race was the most prevalent non-White population, comprising 14.15% of all cases (Table 1). Amongst the cirrhosis cases compared to cases from the general population, cases were more likely to be male (65.67% for the cirrhosis population vs. 59.37% for the general population). For both the HCC general and cirrhosis population, the majority of cases were diagnosed between the ages of 60-70 years (33.82% for the general population and 39.81% for the cirrhosis population). For the HCC general population, the majority of HCC cases were from the Southern and Western regions of the US (34.80% for South and 34.64% for West). HCC cases with liver cirrhosis were more likely to reside in the South and Northeast regions of the US (39.66% for South and 26.60% for Northeast), compared to the cases from the entire general population.

### Model performance

For the general population, LiricNN and LiricLR obtained average AUCs of 0.926 (95% CI: 0.925 to 0.927) and 0.899 (95% CI: 0.898 to 0.900), respectively across nine random runs. We found that LiricNN achieved a higher AUC than LiricLR (*p <* 5*×* 10^*−*189^).

Fig. 2a shows the ROC curves from an arbitrarily chosen run for the general population, where LiricNN and LIRICLR achieved AUCs of 0.925 (95% CI: 0.922 to 0.929) and 0.897 (95% CI: 0.892 to 0.901), respectively.

**Figure 1:**
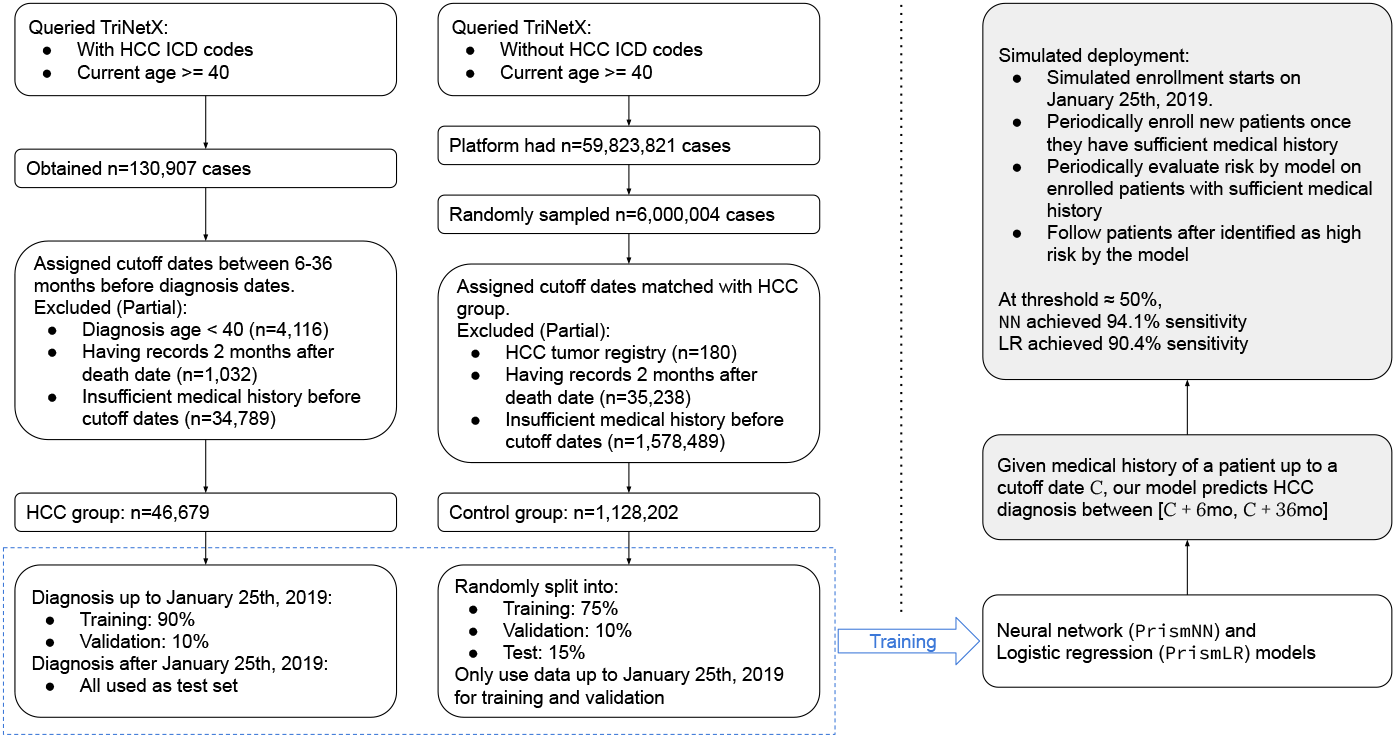
Flowchart outlining key steps of our study

**Figure 2:**
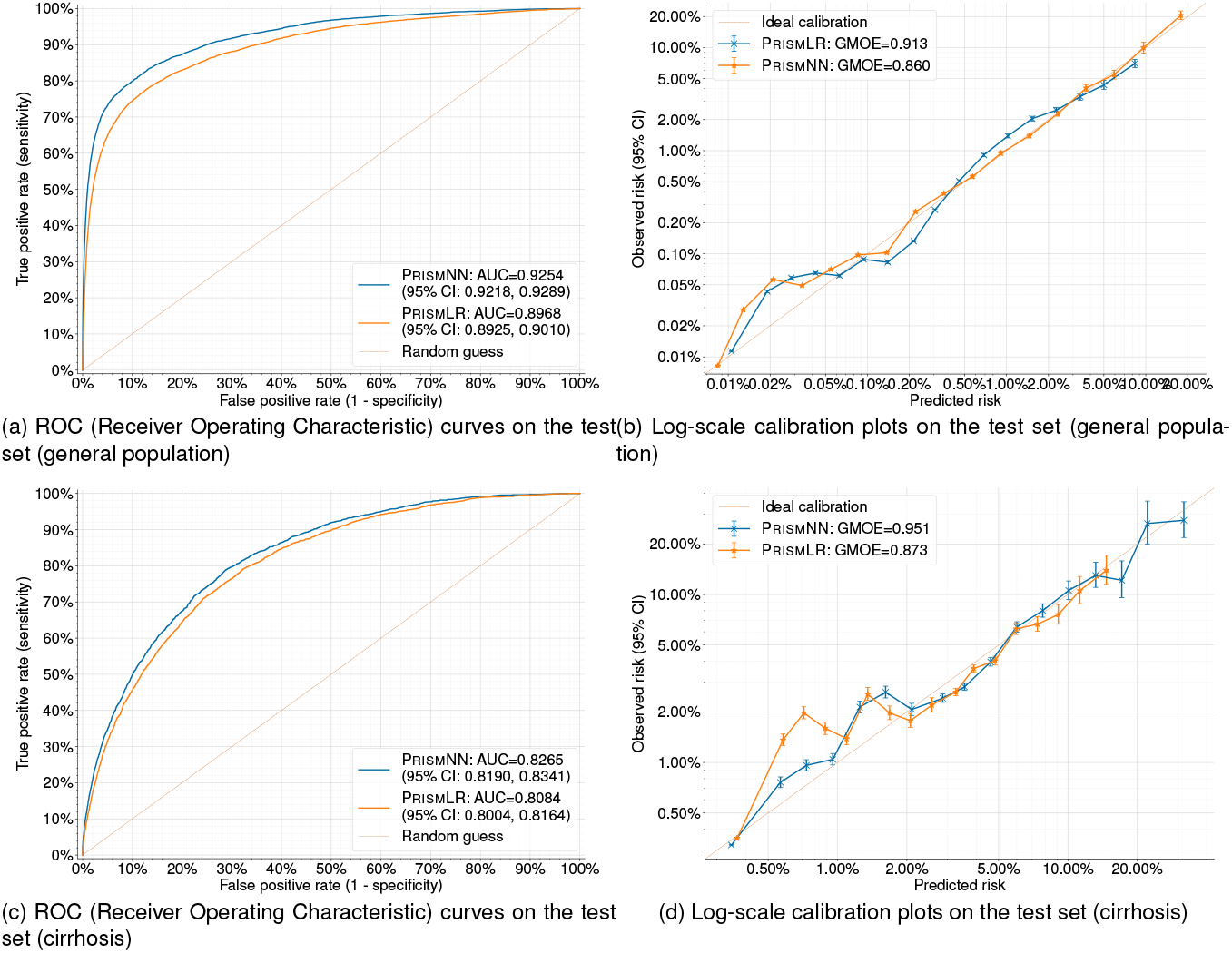
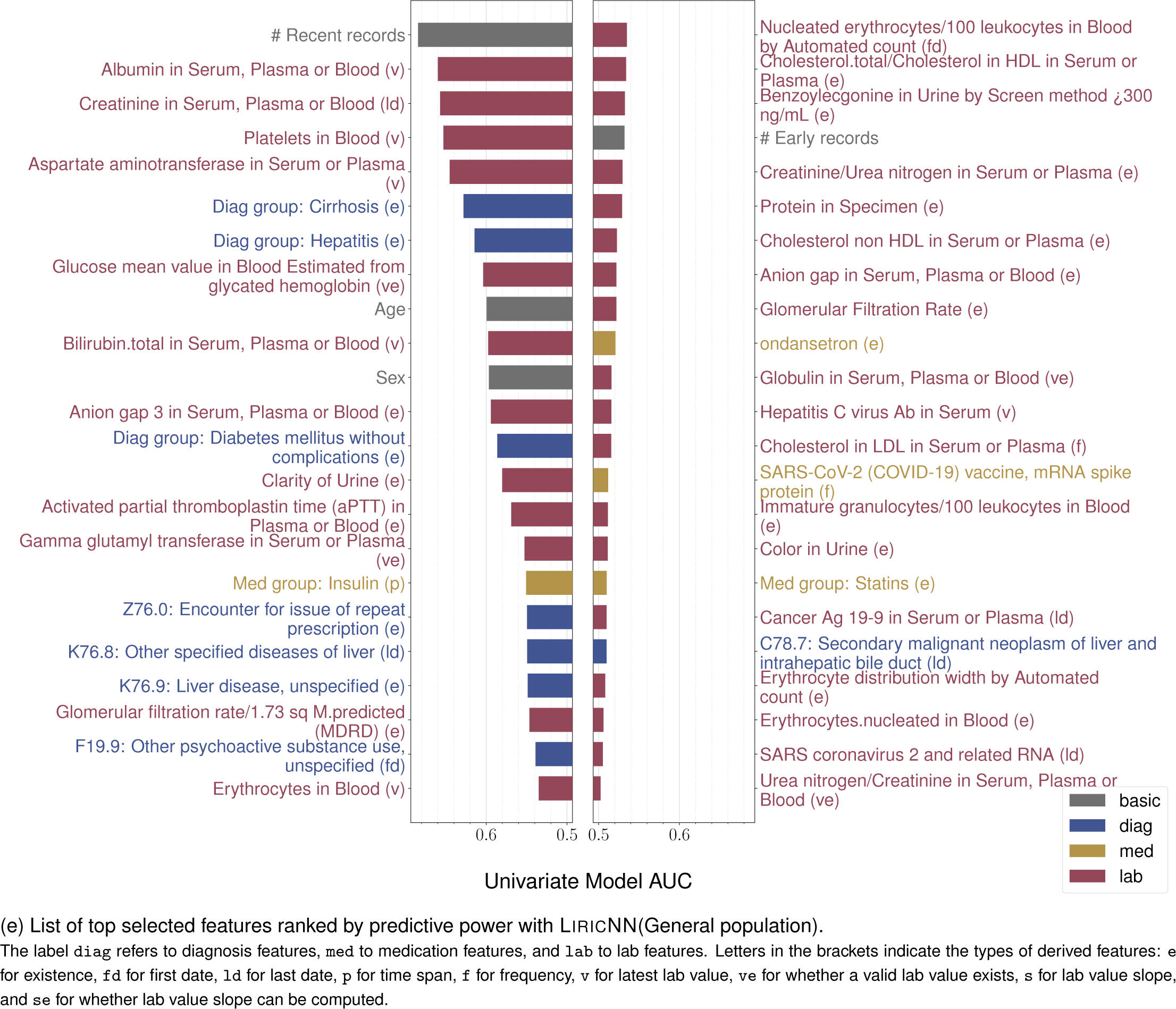
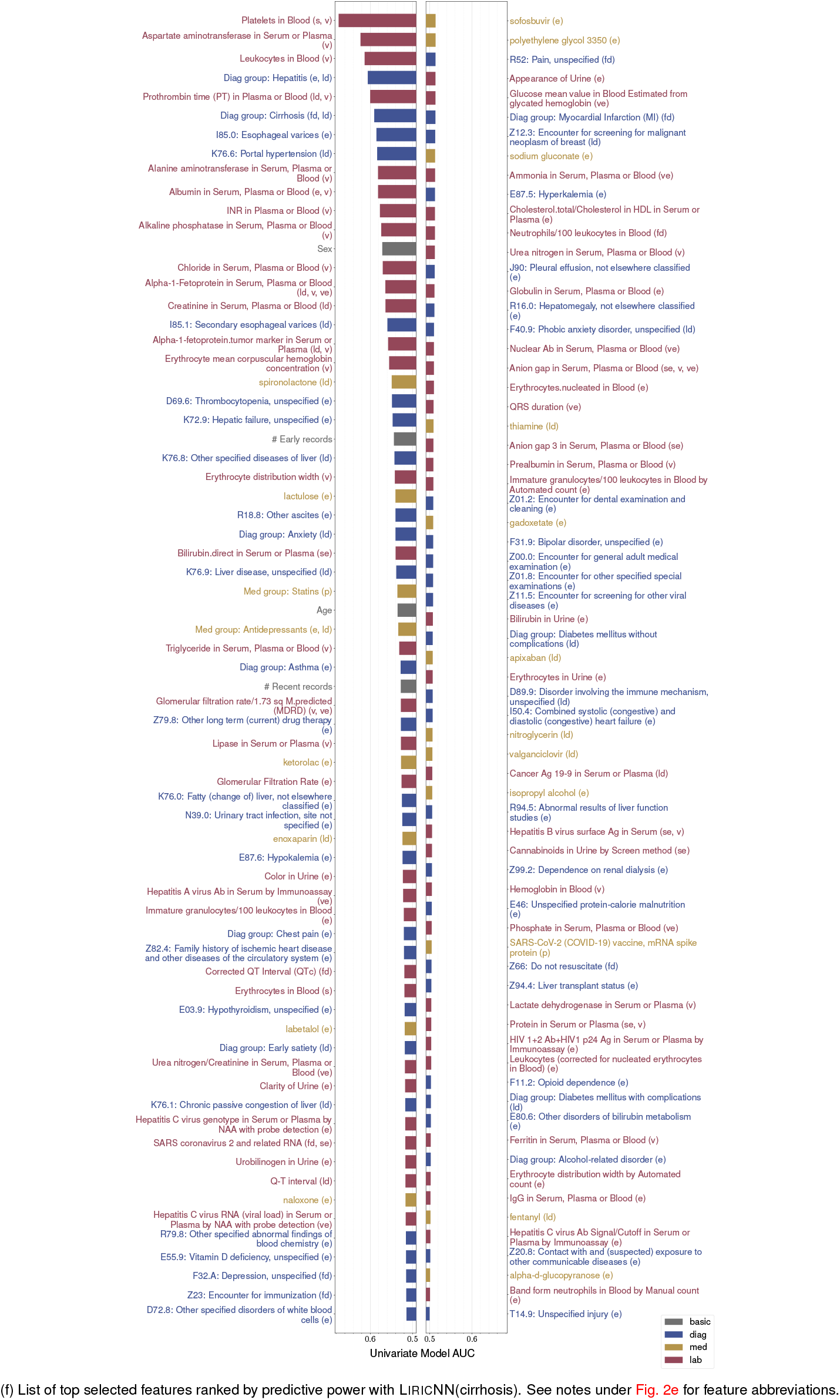
Model evaluation results

Fig. 2b shows the log-scale calibration plots on the test set of the same single run for the general population, where LiricNN and LiricLR scored GMOEs of 0.860 and 0.913. The average GMOEs across nine random runs were 0.887 (95% CI: 0.862 to 0.911) (LiricNN) and 0.969 (95% CI: 0.940 to 0.999) (LiricLR).

For the cirrhosis population, LiricNN and LiricLR obtained average AUCs of 0.829 (95% CI: 0.827 to 0.832) and 0.810 (95% CI: 0.807 to 0.812), respectively across nine random runs. Again, we found that LiricNN achieved a higher AUC than LiricLR (*p <* 10^*−*25^).

Fig. 2c shows the ROC curves from an arbitrarily chosen run for the cirrhosis population, where LiricNN and LiricLR achieved AUCs of 0.827 (95% CI: 0.819 to 0.834) and 0.808 (95% CI: 0.800 to 0.816), respectively.

Fig. 2d shows the log-scale calibration plots on the test set of the same single run for the cirrhosis population, where LiricNN and LiricLR scored GMOEs of 0.951 and 0.873. The average GMOEs across nine random runs were 0.882 (95% CI: 0.841 to 0.923) (LiricNN) and 0.874 (95% CI: 0.826 to 0.923) (LiricLR).

### Feature ranking by predictive power

Fig. 2e presents all the selected features ranked by feature predictive power with LiricNN among the general HCC population. Model features include established features such as those related to diabetes mellitus, cirrhosis, hepatitis, age and sex. Novel features include the number of recent records which is a surrogate for frequency of clinical visits preceding diagnosis, and features related to the lab values of platelets, albumin, aspartate aminotransferase, and bilirubin.

Fig. 2f presents all the selected features ranked by feature predictive power with LiricNN among the cirrhosis HCC population. Model features include those related to the lab values platelets, aspartate aminotransferase, leukocytes, and Prothrombin Time, as well as the date of cirrhosis and hepatitis diagnosis, esophageal varices, portal hypertension, age and sex.

### Internal-external validation results

Fig. 3 and Fig. 4 shows results for location-based, race-based, and temporal external validations for the general HCC and cirrhosis HCC populations respectively.

**Figure 3:**
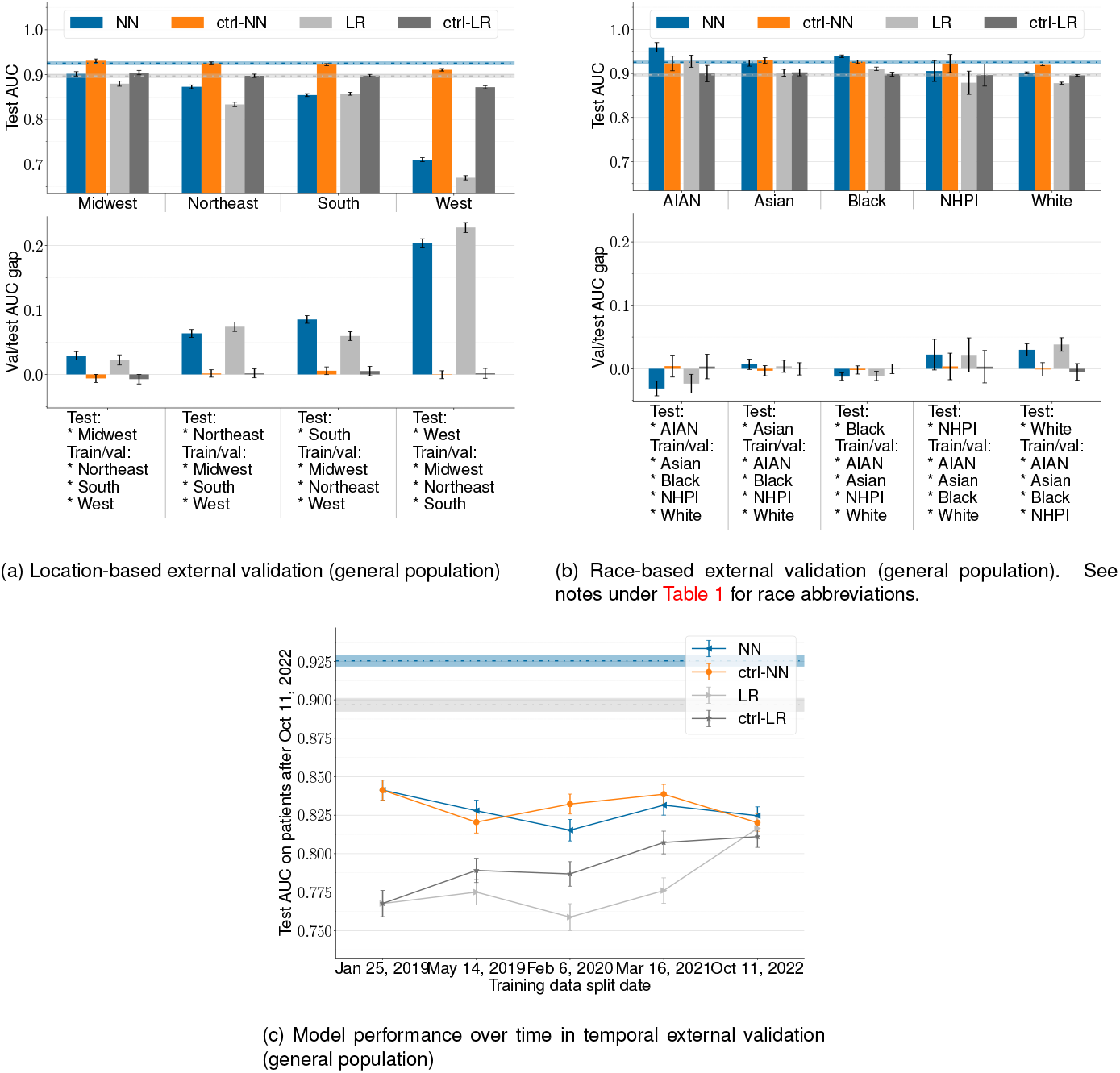
Results for location-based, race-based, and temporal external validations (general population). Error bars indicate 95% CI. <monospace>NN</monospace> is short for LiricNN, <monospace>LR</monospace> short L<monospace>IRIC</monospace>LR, and <monospace>ctrl-</monospace> for control models with matched training data size.

**Figure 4:**
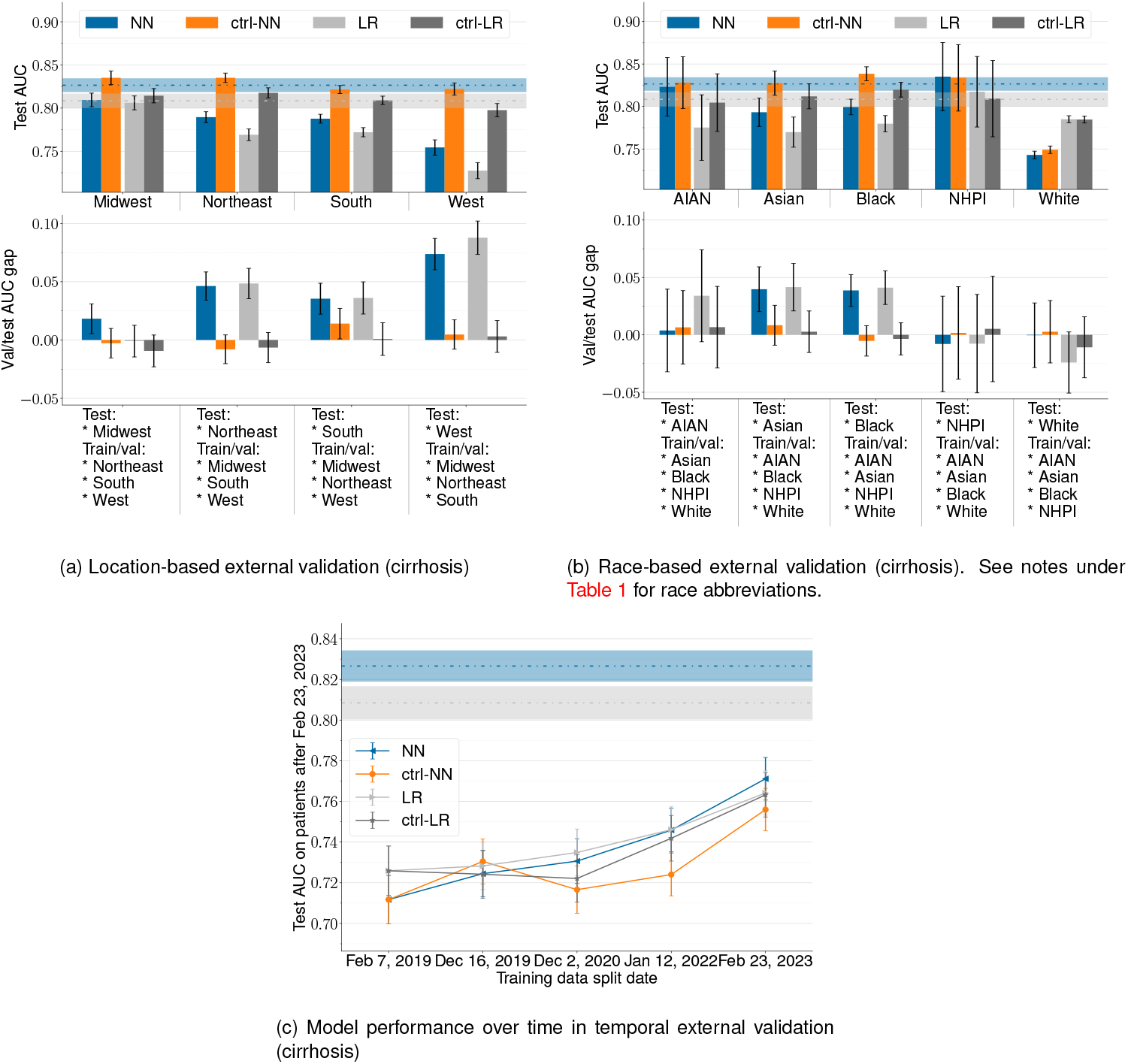
Results for location-based, race-based, and temporal external validations (cirrhosis). Error bars indicate 95% CI. <monospace>NN</monospace> is short for L<monospace>IRIC</monospace>NN, <monospace>LR</monospace> short LiricLR, and <monospace>ctrl-</monospace> for control models with matched training data size.

For the general population, the models showed similar performance across all racial groups and various geographic locations, except for the West where performance was diminished.

For the Midwest, Northeast, South, and West regions, LiricNN test AUCs were 0.902 (95% CI: 0.897 to 0.906), 0.873 (95% CI: 0.868 to 0.877), 0.854 (95% CI: 0.851 to 0.857), and 0.710 (95% CI: 0.706 to 0.714) (*average*: 0.835 (95% CI: 0.690 to 0.980)). Slightly lower, LiricLR test AUCs were 0.880 (95% CI: 0.874 to 0.885), 0.833 (95% CI: 0.828 to 0.838), 0.857 (95% CI: 0.854 to 0.860), and 0.669 (95% CI: 0.665 to 0.674) (*average*: 0.810 (95% CI: 0.648 to 0.972)).

These were 0.029 to 0.201 lower than the respective control models for LiricNN and 0.025 to 0.201 for LiricLR.

For the AIAN, Asian, Black, NHPI, and White racial groups, LiricNN test AUCs were 0.959 (95% CI: 0.949 to 0.970), 0.923 (95% CI: 0.917 to 0.930), 0.939 (95% CI: 0.936 to 0.942), 0.906 (95% CI: 0.882 to 0.929), and 0.902 (95% CI: 0.900 to 0.904) (*average*: 0.926 (95% CI: 0.882 to 0.969)).

Slightly lower, LiricLR test AUCs were 0.899 (95% CI: 0.859 to 0.939) (*average*: 0.899 (95% CI: 0.859 to 0.939)). This performance has a statistically insignificant difference compared to the respective control models (*−*0.037 to 0.018 for LiricNN and *−*0.028 to 0.018 for LiricLR).

Fig. 3c demonstrates temporal validation results. LIRICNN and LiricLR achieved average test AUCs of 0.828 (95% CI: 0.810 to 0.846) and 0.779 (95% CI: 0.739 to 0.819), respectively.

For the cirrhosis population, the models showed similar performance across different geographic locations and racial groups.

For the Midwest, Northeast, South, and West regions, LiricNN test AUCs were 0.809 (95% CI: 0.802 to 0.817), 0.790 (95% CI: 0.783 to 0.796), 0.788 (95% CI: 0.783 to 0.793), and 0.754 (95% CI: 0.746 to 0.763) (*average*: 0.785 (95% CI: 0.746 to 0.825)). Slightly lower, LiricLR test AUCs were 0.806 (95% CI: 0.798 to 0.814), 0.769 (95% CI: 0.762 to 0.776), 0.772 (95% CI: 0.767 to 0.777), and 0.728 (95% CI: 0.718 to 0.737) (*average*: 0.769 (95% CI: 0.714 to 0.824)).

These were 0.026 to 0.068 lower than the respective control models for LiricNN and 0.008 to 0.068 for LiricLR.

For the AIAN, Asian, Black, NHPI, and White racial groups, LiricNN test AUCs were 0.823 (95% CI: 0.789 to 0.858), 0.793 (95% CI: 0.776 to 0.810), 0.799 (95% CI: 0.790 to 0.809), 0.835 (95% CI: 0.795 to 0.875), and 0.743 (95% CI: 0.738 to 0.747) (*average*: 0.799 (95% CI: 0.731 to 0.866)).

Slightly lower, LiricLR test AUCs were 0.785 (95% CI: 0.743 to 0.828) (*average*: 0.785 (95% CI: 0.743 to 0.828)). This performance has a statistically insignificant difference compared to the respective control models ( −0.001 to 0.039 for LiricNN and 0.008 to 0.042 for LiricLR).

Fig. 4c demonstrates temporal validation results. LIRICNN and LiricLR achieved average test AUCs of 0.737 (95% CI: 0.695 to 0.778) and 0.740 (95% CI: 0.710 to 0.770), respectively.

### Simulated deployment results

For the general population, we simulated model deployment with enrollment from Feb 7, 2020 to Nov 3, 2020 on 145,875 patients (including 8,660 HCC cases) in the test set. Mean ages at enrollment and at HCC diagnosis were 61.36 (SD 11.64) and 67.32 (SD 9.53). Patients were followed up for 3.14 (SD 0.23) years on average. Table 2 shows sensitivity, specificity, PPV estimation on the entire TriNetX population, MEPD, NNS, No. in bin, and No. of cases metrics achieved given different risk levels predicted by LiricNN and LIRICLR.

For the cirrhosis population, we simulated model deployment with enrollment from Dec 3, 2020 to Mar 3, 2021 on 27,539 patients (including 3,426 HCC cases) in the test set. Mean ages at enrollment and at HCC diagnosis were 62.43 (SD 10.28) and 66.41 (SD 8.99). Patients were followed up for 2.74 (SD 0.06) years on average. Table 3 shows sensitivity, specificity, PPV, MEPD, NNS, No. in bin, and No. of cases metrics achieved given different risk levels predicted by LiricNN and LiricLR.

## Discussion

We developed, validated, and simulated the deployment of LIver RIsk Computation (Liric) models. Liric models can accurately identify individuals at high-risk of HCC from the general population, three years before diagnosis, using routinely collected patient clinical and lab data from 64 HCOs across the US. Models were trained on 46,679 HCC cases and 1,128,202 controls. LiricNN achieved an AUC of 0.93 and LiricLR an AUC of 0.90 on the test sets. Calibration measuring the predicted risks to the actual risks was excellent, with a GMOE of 0.86 for PrismNN and 0.91 for PrismLR. During training, features were automatically derived from demographics, diagnoses, medications, and lab results using a data-driven feature selection approach. Typical AI models operate opaquely, where the specific features used by a model or the decision process based on those features are not understood by humans. However, we prioritized enhanced interpretability and evaluated which features were top predictors for the model. This process identified 46 features, which encompass previously recognized factors associated with HCC development such as age, sex, cirrhosis, hepatitis, and diabetes mellitus. Additionally, it facilitated the discovery of novel features influencing model predictions, such as the frequency of clinical encounters prior to an HCC diagnosis, as well as serum levels of platelets, albumin, and aminotransferase values.

Another issue for AI-derived models is the concern for racial bias and healthcare disparities [34]. To address this concern, we evaluated our models across different races and geographic locations across the country. Race-based internal-external validation demonstrated model generalizability across diverse populations with minimal performance drop. Results of location-based validation showed modest AUC drops, implying that there are systematic differences between EHR data from geographically different HCOs. Models maintained good performance in temporal validation.

To assess model performance in a clinical setting, we simulated model deployment on data from approximately 145,000 patients over 3 years, employing a prospective cohort design. The simulation produced a table with multiple risk thresholds, facilitating the categorization of individuals into low, intermediate, and high-risk groups. Risk groupings optimize screening strategies by omitting low-risk individuals from screening and intensifying efforts to target those at high risk. When designing a potential screening program for a relatively rare disease like HCC, it is critical to define an at-risk population, such that the prevalence warrants screening. The approach described in this report does not define risk groups per se but demonstrated how the models can be used to define appropriate risk stratification based on various cut-offs. We emphasize the need for engagement with multiple stakeholders to strike a balance between patient benefits, harms, and cost-effectiveness in determining risk group cut-offs, which is beyond the scope of this paper.

AI predictive models have the potential to greatly impact HCC screening strategies by concentrating early detection efforts on a specific population with the highest risk. Additionally, these models can identify individuals who are at risk but currently ineligible for screening, such as those with Nonalcoholic fatty liver disease (NAFLD) [35, 36, 13]. Risk-stratified screening has been proven to be more costeffective than the current HCC screening guidelines [37]. Moreover, these efforts could enhance the utilization of established screening methods like abdominal sonography and alpha-fetoprotein [38] and would enable the allocation of crucial resources to those at highest risk. However, existing AI models for HCC face limitations due to their restricted sample sizes and lack of diversity, as well as their accuracy, generalizability, and interpretability [39]. Furthermore, these models largely lack a comprehensive plan for future implementation into the clinical workflow [40].

We defined the following goals for our models: (1) focusing screening efforts on a more targeted population to enhance the utilization of existing screening modalities and (2) effectively identifying individuals often overlooked by conventional guidelines, for example, those without underlying cirrhosis. Our work encompasses a comprehensive range of stages involved in risk prediction modeling [41]. This includes delineating clinical quality improvement objectives, developing the models, conducting both internal and external validation, and assessing potential biases. Furthermore, our efforts extend to the meticulous preparation required for clinical deployment, with the ultimate aim of seamlessly integrating the models into the clinical workflow. To streamline this process, we utilized a federated EHR network, providing us with the framework to carry out each of these steps. The federated network platform has enabled us to successfully overcome common challenges associated with utilizing data from multiple institutions that operate on different EHR systems. This includes streamlining processes like data aggregation and standardization. Within this network, de-identified data from diverse EHR systems is securely collected, and virtually consolidated from all participating Healthcare Organizations (HCOs). The data obtained from various EHR systems undergoes standardization and harmonization, ensuring a consistent dataset for model development. This platform also addresses concerns related to confidentiality, as all data is deidentified, safeguarding the privacy of patients and participating healthcare institutions.

### Overfitting, external validation, generalizability

Our models were trained and externally validated on 46,679 HCC cases and 1,128,202 controls from academic medical centers, community hospitals, and outpatient clinics from 64 HCOs across the USA. By utilizing a large and heterogenous population, including individuals from various ethnic backgrounds spanning different geographic locations for training, we ensured the development of robust and generalizable models while mitigating the risk of overfitting to the training population. Models were trained on the entire population and rigorously tested on each race and geographic location independently. The outcomes of our external validation revealed marginal disparities in performance, indicating strong alignment between the test model and the external validation results (Figs. 3 and 4).

These results strongly suggest that Liric models can be effectively applied to new populations nationwide, yielding comparable outcomes. The transferability of this model to diverse populations holds significant implications for identifying high-risk racial and ethnic groups in specific geographic areas. This transferability also extends to resource allocation for HCC screening education targeted towards patients and caregivers, which has been proven effective in increasing utilization of screening and lowering mortality rates in specific populations [38].

### Interpretability

While employing a predefined set of features known to be correlated with HCC [21, 22, 24] facilitates the creation of easily explainable models, this approach limits the potential for discovering novel features that could significantly enhance predictive performance and drive future causal studies. Neural networks, in particular, are often perceived as “black box” models due to the inherent challenge of comprehending their internal workings. This is a consequence of the complex non-linear relationships between features utilized by the model to generate predictions. To enhance the interpretability of our models, we have developed a highly efficient algorithm for automated feature selection tailored specifically for large-scale data in neural networks (Ref- Kai Jia). This algorithm equips us with the means to uncover and utilize the most informative features while preserving the interpretability of the model.

Through the utilization of our feature selection algorithm, we have successfully XXX identified XXX the 46 features employed by LiricNN to compute the risk of HCC, enabling us to establish a solid basis for interpreting these models. Additionally, we have developed a model tailored to patients with liver cirrhosis, the demographic currently undergoing HCC screening, for comparison. While both models utilize features associated with platelet, albumin, and aminotransferase values, the general population model also utilizes diabetes-related features, including the diabetes mellitus code, time span of insulin use, and glucose values. Reliance on diabetes features, which have been linked to NAFLD, and represents the most prevalent cause of HCC development in non-cirrhotic livers [42], may suggest that the model possesses the capability to effectively identify HCC cases that arise even in the absence of underlying cirrhosis. In contrast, the cirrhosis model utilizes features related to complications of advanced liver disease, such as portal hypertension, primary and secondary esophageal varices. Both models rely on the patient having a cirrhosis or hepatitis diagnosis, but while the general population model relies on the existence of these diagnostic codes, the cirrhosis model uses the date of their appearance, or their timing relative to HCC diagnosis, as a top feature. The comparison between the two models suggests that the general population model encompasses a broader at-risk population for HCC, extending beyond the currently screened cirrhosis population.

Notably, LiricNN incorporates novel features, such as “number of recent records” and “number of early records.” The former refers to the count of diagnosis, medication, or laboratory entries in the EHR within 36 months preceding the cutoff date, while the latter represents the count of such entries more than 36 months prior to the cutoff date. These features serve as proxies for the frequency of clinical encounters leading up to the HCC diagnosis. Increased clinical encounter frequency prior to cancer diagnosis has previously been correlated with the likelihood of cancer detection [43]. However, this aspect has not been previously utilized in HCC risk prediction models until now.

Overall, the inclusion of these diverse and novel features in Liric models enhances their ability to capture and predict HCC cases, including those arising in patients with noncirrhotic livers.

Our current comprehension of each feature’s trend or its exact impact mechanism on the risk score is limited. Derived features may have both positive and negative effects on risk scores, and we lack a comprehensive understanding of each feature in isolation. Instead, we perceive these features as interacting collectively and non-linearly to make predictions. For example, statins, part of the model’s selected features, are known to be protective factors for HCC [44], whereas diabetes has been previously correlated with increased HCC risk [45].

### Missing Data

When utilizing Electronic Health Record (EHR) data for modeling, a well-known challenge is the occurrence of “missing data.” We do not view missing data in the EHR as a random process, but recognize it as a reflection of the impact of the healthcare system on data quality [29], as well as an inherent characteristic of the EHR itself. For instance, one of the predictive features in our model is the frequency of clinical visits prior to HCC diagnosis. This increase in physician office visits, often driven by new or recurring symptoms, has previously shown correlation with an impending cancer diagnosis [43], a relationship we anticipate holds true in our study as well.

Based on this understanding of the nature of EHR we have adopted a specific approach to address missing data. We incorporate an existence bit feature that indicates the availability of a particular EHR entry type in a patient’s medical record. This encoding enables our model to make predictions based on the presence or absence of a feature. We did not use data imputation since LiricNN uses sophisticated nonlinear reasoning, and imputation provides little to no useful additional information for these models.

### Conclusion

In conclusion, we have successfully developed, validated, and simulated the deployment of Liric, liver cancer risk prediction models that exhibits the capability to identify individuals at high risk of developing HCC 6-36 months prior to diagnosis. The external validation results affirm the generalizability and transferability of the Liric models across a diverse patient population nationwide, potentially bridging the social and geographic disparities associated with specialized care predominantly accessible in affluent urban areas. By increasing awareness among primary care physicians (PCPs) and facilitating the identification of high-risk individuals from the general population, Liric models have the potential to enhance HCC screening efforts by leveraging existing modalities. Furthermore, the models focus on the highest-risk cirrhotic patients and can identify individuals who require screening despite the absence of underlying cirrhosis.

Liric models, which utilize routinely collected EHR data, can potentially be integrated into EHR systems so that primary care providers are alerted to the patient’s risk of HCC, allowing for timely actions to reduce the likelihood of missed diagnoses. The interpretability of Liric models can enhance trust among physicians, which is a crucial factor for their future implementation.

To assess the real-time performance and validate the model’s efficacy, a prospective validation study is warranted. This study would offer crucial insights regarding the model’s reliability and utility in clinical practice, further solidifying its role in HCC prediction and enhancing patient outcomes.

## Data Availability

The de-identified data in TriNetX federated network database can only be accessed by researchers that are either part of the network or have a collaboration agreement with TriNetX. As stated in the manuscript, we accessed data as part of a no-cost collaboration agreement between BIDMC, MIT, and TriNetX.

## Abbreviations

EHR: electronic health record
HCC: Hepatocellular Carcinoma
GP: General Population
NAFLD: non-alcoholic fatty liver disease
HCO: health care organization
LR: logistic regression
NN: neural network
AI: Artificial Intelligence

## Acknowledgements

The authors are grateful to Gadi Lachman and TriNetX for providing support and resources for this work. MR, LA, KJ, BG, PS acknowledge the contribution of resources by TriNetX, including the provision of secured laptop computers, access to the TriNetX EHR database, and clinical, technical, legal, and administrative assistance from their team of clinical informaticists, engineers, and technical staff. LA acknowledges support from Coverys Community Healthcare Foundation for this work. MR and KJ received funding from DARPA and Boeing. MR also received funding from the NSF and Aarno Labs. SK, MP, and JW are employees of TriNetX.

## Declaration of AI and AI-assisted technologies in the writing process

During the preparation of this work the author(s) used ChatGPT in order to enhance the writing style and improve the readability of this manuscript. After using this tool, the author(s) reviewed and edited the content as needed and take(s) full responsibility for the content of the publication.

